# The global burden of chikungunya virus and the potential benefit of vaccines

**DOI:** 10.1101/2024.10.24.24315872

**Authors:** Gabriel Ribeiro dos Santos, Fariha Jawed, Christinah Mukandavire, Arminder Deol, Danny Scarponi, Leonard E.G. Mboera, Eric Seruyange, Mathieu J.P. Poirier, Samuel Bosomprah, Augustine O. Udeze, Koussay Dellagi, Nathanael Hozé, Jaffu Chilongola, Gheyath K. Nasrallah, Elmar Saathof, Simon Cauchemez, Henrik Salje

## Abstract

The first chikungunya virus (CHIKV) vaccine has now been licensed, however, its potential to reduce disease burden remains unknown due to a poor knowledge of the underlying global burden. We use data from seroprevalence studies, observed cases and mosquito distributions to quantify the underlying burden in 190 countries and territories, and explore the potential impact of the vaccine. We estimate that 104 countries have experienced transmission, covering 2.8 billion individuals and that in epidemic settings, the mean duration between outbreaks is 6.2 years, with 8.4% of the susceptible population infected per outbreak. Globally there are 33.7 million annual infections, driven by countries in Southeast Asia, Africa and the Americas. Assuming a vaccine efficacy against disease of 70% a protection against infection of 40%, vaccinating 50% of individuals over 12 years old in places and times where the virus circulates would avert 3,718 infections, 2.8 deaths and 158 DALYs per 100,000 doses used. These findings highlight the global burden and the significant potential of the vaccine.

## Introduction

Chikungunya virus (CHIKV) is an alphavirus transmitted by *Aedes* mosquitoes. Infection in humans is characterised by acute symptoms of rash and fever. In addition, there is frequently severe joint pain that can last for many months^1^. Around one in a thousand cases result in death, mainly in the elderly^2^. Cases of chikungunya have been found throughout tropical and subtropical countries around the world^3^. In many places, CHIKV transmission consists of outbreaks, followed by periods without circulation. However, endemic transmission has also been reported^4–6^. Following decades with few effective tools to combat CHIKV, significant investment by the Coalition of Epidemic Preparedness Innovations (CEPI) has led to the licensure by the Food and Drug Administration (FDA) of the first CHIKV vaccine, IXCHIQ (VLA1553), developed by Valneva^7^. Due to the unpredictable nature of CHIKV epidemiology, licensure was obtained through a correlate of protection rather than traditional phase III trials.

A major hurdle in the optimal deployment of the vaccine is the limited understanding of the underlying burden from CHIKV around the globe, and in addition how best to use the vaccine. The decision whether to use a vaccine typically relies on a vaccine investment case, which quantifies the impact of using a vaccine on the number of infections, cases and deaths averted. However, in the case of CHIKV, we have a very poor understanding on where the virus circulates, hampering the development of investment cases. The Gavi Alliance which helps Lower Income countries purchase vaccines has placed CHIKV vaccines on a Learning Agenda, which means that it does not feel there is sufficient information available to make informed decisions on the likely impact of the vaccine^8^. This knowledge gap is driven by frequent clinical misdiagnosis with other pathogens such as dengue or influenza, and limited access to confirmatory testing^9^. Further, it is unclear whether the epidemic nature of the virus means that the vaccine could be deployed from stockpiles in response to detected outbreaks, rather than the integration into immunisation schedules.

In order to explore the potential of the vaccine on the global burden of CHIKV, we can combine the results of case reports, seroprevalence studies that identified antibodies to CHIKV and mathematical modelling to estimate the underlying number of annual infections per country^10^. We can then use a transmission model to critically assess the potential impact of vaccination campaigns to avert future infections, cases and deaths, providing an evidence base to guide future vaccine use.

## Results

We conducted a literature review to identify whether individual countries and territories have ever reported evidence of local CHIKV transmission. We considered all countries and territories (later referred as countries) that have a population size of over 200,000 individuals (N=180, UN population size estimates). We found CHIKV had been directly reported in 91 countries and territories, representing 51% of all locations (Figure 1A). We found that whether or not a country had ever reported CHIKV transmission was strongly correlated with the population-weighted estimated presence of *Aedes aegypti* (Pearson correlation: 0.89) and *Aedes albopictus* (Pearson correlation: 0.82) in the country (Figure 1B) (1, 2).

**Figure 1.**
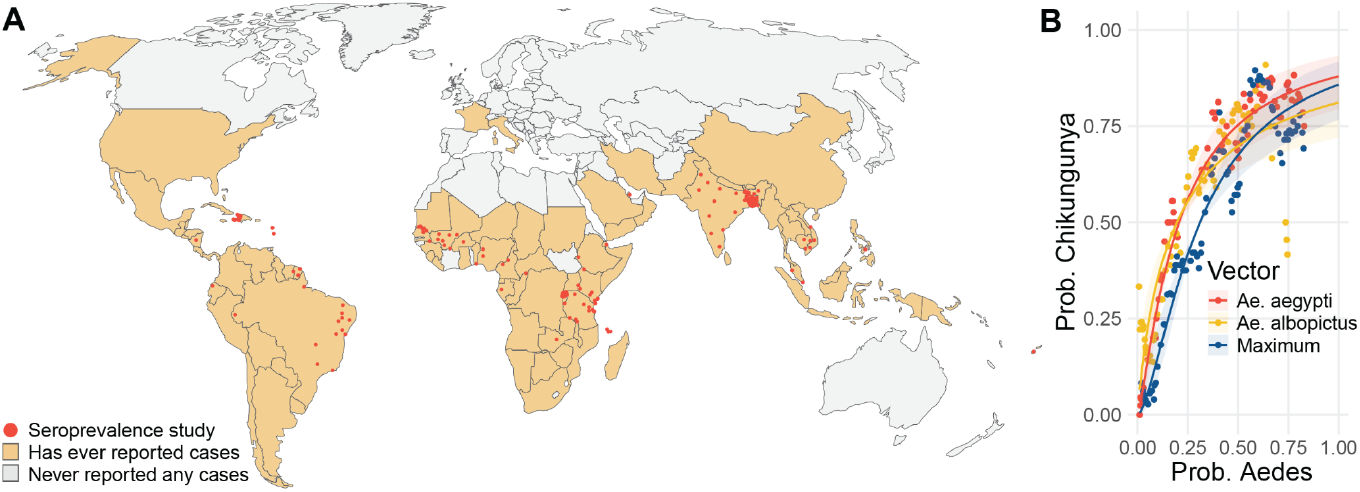
**(A)** Countries with a history of CHIKV transmission and location of seroprevalence studies **(B)** Relationship between average estimated occurrence of *Aedes aegypti* and *Aedes albopictus* in a country with the probability that CHIKV has ever been reported in that country. The probability of Aedes occurrence was calculated using existing estimates of mosquito distributions in 5km x 5km grid cells throughout the globe ^22^. For each country we calculated the human population weighted average occurrence in the country (so i.e., mosquito levels in urban hubs provided more weight than rural locations). The plot shows among groups of countries with a similar level of Aedes occurrence, the proportion of countries that had ever reported CHIKV outbreaks. We separately consider *Aedes aegypti, Aedes albopictus* and a ‘Maximum’, which is the maximum of either *Aedes aegypti* or *Aedes albopictus* in any location. The lines present the fit of a logistic regression model.

We next categorised all countries into ‘epidemic’ (i.e., evidence of transmission but not sustained across years), ‘endemic’ (i.e., with evidence of sustained transmission each year), and ‘no transmission’ based on the presence of detected cases, seroprevalence studies and the presence of the mosquito vector. We also generated an assessment of the strength of the evidence for the categorization of each country (Table 1). Overall, we find evidence of endemic transmission in 6 countries, 4 in East Africa (Kenya, Mozambique, Rwanda and Tanzania), 1 located in South America (Brazil), and 1 in Asia (India). We find evidence of epidemic transmission in a further 98 countries (Figure 2, Table S1). Of these, 13 have never reported any cases but they have high levels of the vectors and neighbouring countries where transmission has been reported. We find no evidence of transmission in a further 76 countries.

**Table 1.** Criteria used to define epidemic status by country. Confirmed cases are those that are either PCR or IgM confirmed.

| Epidemic status | Criteria |
| --- | --- |
| Endemic - Good evidence | Confirmed autochthonous cases each year for the past 5 years. In the Americas, which has better disease surveillance, we include a criteria for a minimum of 1,000 detected autochthonous cases per year. |
| Endemic - Poor evidence | Confirmed autochthonous cases each year in at least 3 of the past 5 years and/or serological evidence of endemic transmission. In the Americas, which has better disease surveillance, we include a criteria for a minimum of 100 detected cases per year. |
| Epidemic - Good evidence | Confirmed cases since 2010<br>OR<br>Serological evidence of transmission since 2010 |
| Epidemic - Poor evidence | Confirmed cases prior to 2010<br>OR<br>serological evidence of transmission prior to 2010<br>OR<br>In low HAQ countries, no confirmed cases but probability *albopictus/aegypti* mosquitoes present >0.5 |
| No transmission - Good evidence | In high HAQ countries, no confirmed cases<br>OR<br>Everywhere else, no confirmed cases and probability *albopictus/aegypti* mosquitoes present <0.1 |
| No transmission - Poor evidence | No confirmed cases and probability *albopictus/aegypti* mosquitoes present 0.1-0.5 |

**Figure 2.**
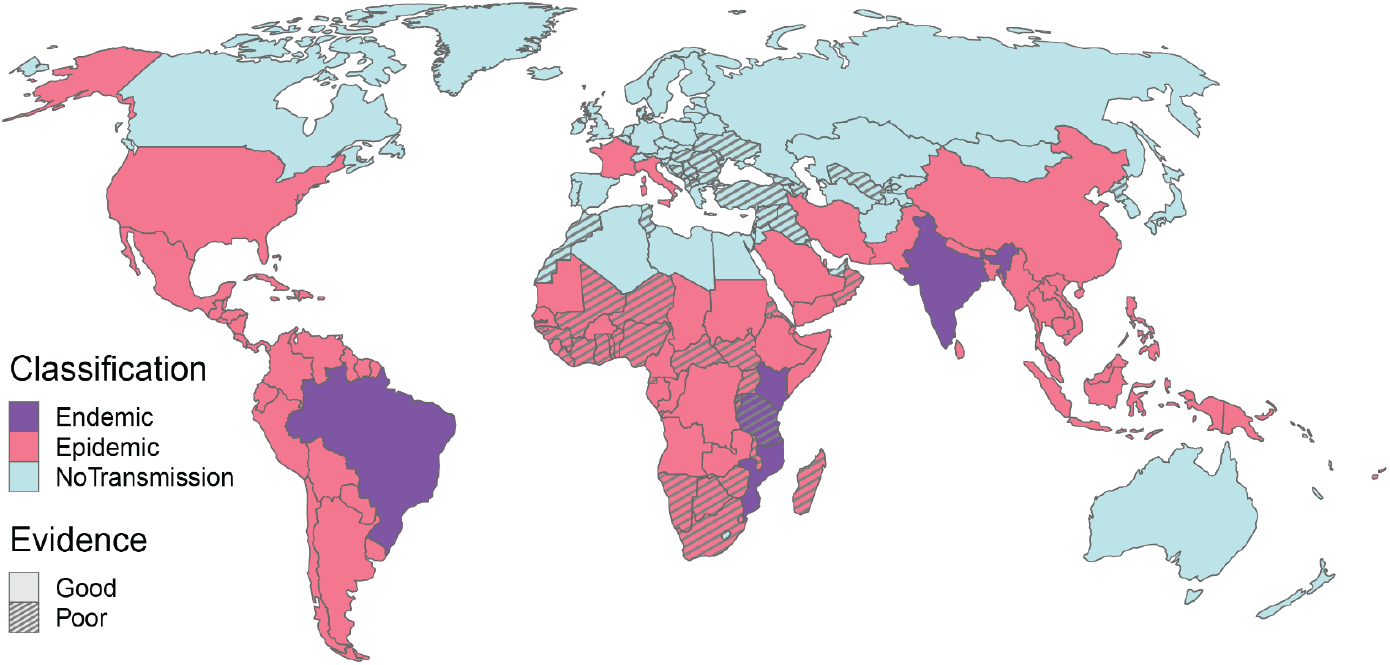
Map of endemic and epidemic countries and level of evidence.

To obtain an estimate of the size of the population in each country that is at risk of CHIKV infection we used our estimated association between the occurrence of mosquitoes (defined here as the greater of either *Aedes aegypti* or *Aedes albopictus* in each location) and the national probability of CHIKV transmission (Figure 1B). For each 5 km x 5 km grid cell, we used the relationship between mosquito occurrence and the probability of CHIKV outbreaks and the number of people living in that cell to obtain a weighted average of the size of the population at risk in that location. For India, China and the United States that have large populations, we introduce an additional mask that assumes CHIKV transmission only occurs in areas where there is good evidence of sustained transmission, from either national seroprevalence studies (India), or good case reporting (China and the United States)^6^. This approach results in an average of 68% of the population in endemic and epidemic countries being at risk of CHIKV infection (Figure S1). Overall, we estimate that 2.8 billion individuals globally live in locations at risk of CHIKV transmission.

We next fit serocatalytic models to the results of 48 age-specific seroprevalence studies collected from 29 countries to estimate the average dynamics of CHIKV transmission in endemic and epidemic countries (Figure S2). We find that in endemic locations, the mean annual probability of becoming infected among the susceptible population was 2.4% (95%CI: 1.8%-3.5%), ranging from 0.17% to 7.4% across the 24 different endemic locations in our datasets. In epidemic settings, the mean annual probability of infection was 1.6% (95%CI: 1.3%-2.4%), ranging from 0.04% to 6.5% across locations. However, we find that on average, there is no transmission in most years, with an annual probability of an outbreak of 15.9% (95%CI 13.1%-19.6%), equivalent to a duration of 6.2 years between outbreaks (95%CI: 5.1-7.6). We estimated the mean proportion of the susceptible population that gets infected during outbreaks was 8.4% (95%CI: 7.2%-9.1%).

Overall, these estimates allow us to reconstruct the average number of infections occurring annually within each country. We estimate that globally, there are 33,700,000 infections per year (95%CI: 19,800,000-55,600,000). The most affected WHO region is Southeast Asia, followed by Africa and the Americas (Figure 3A, Figure S3, Table S2). We estimate there are 13,800,000 infections in endemic countries, driven largely by India (9,100,000 annual infections) and 19,900,000 infections in epidemic countries. Given standard assumptions of 50% symptomaticity rates and 20% probability of an infection leading to chronic sequelae, we estimate that these infections lead to 16,900,000 symptomatic cases, of which 3,360,000 result in chronic sequelae and 22,600 deaths (95%CI: 13,300-36,900)^10^. Overall, we estimate there are 1,070,000 DALYs lost to CHIKV each year (Figure 3B).

**Figure 3.**
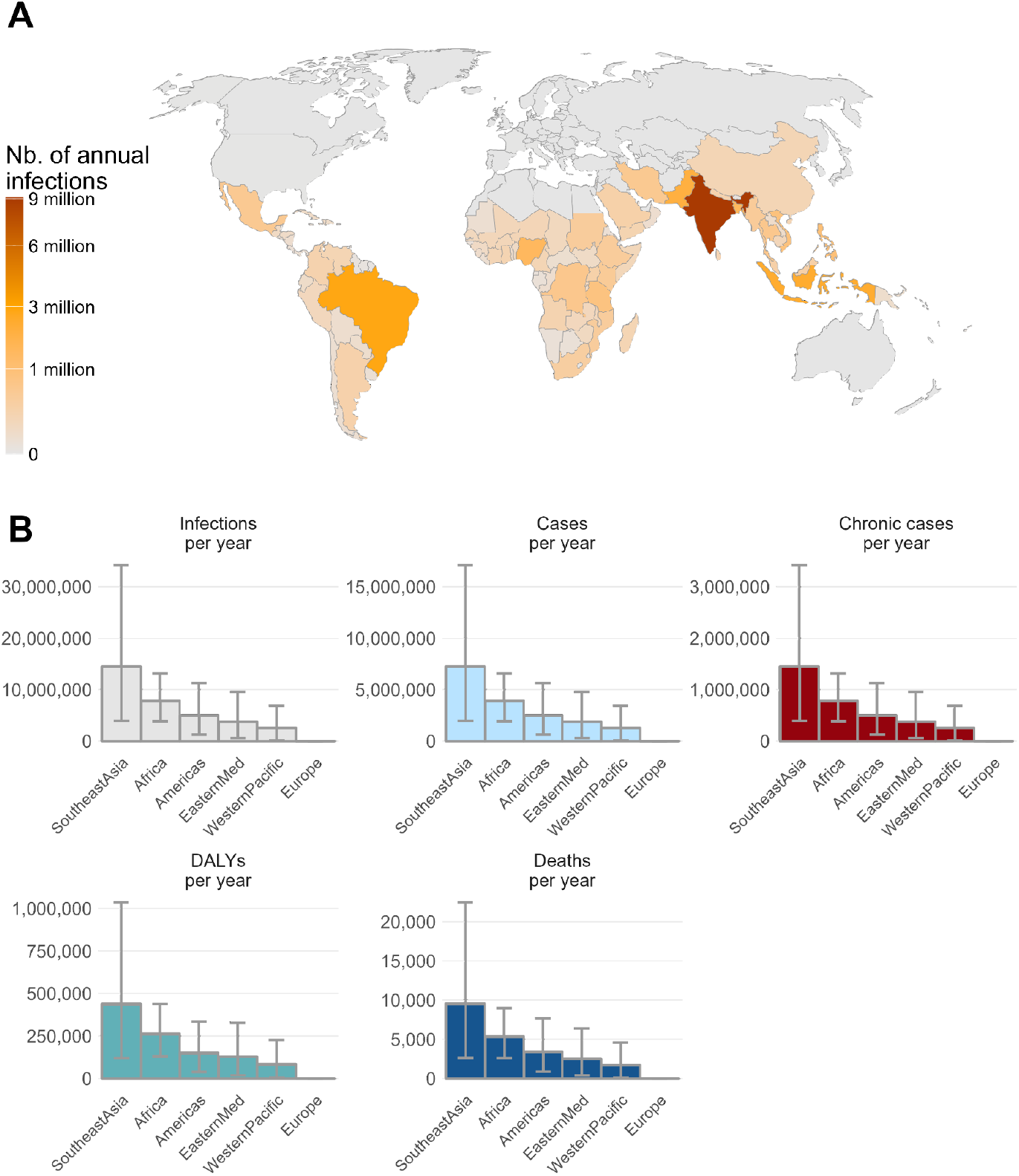
**(A)** Annual infections per country **(B)** Annual infections, cases, chronic cases, DALYs and deaths per WHO region

Using our estimates of the underlying level of infection, we estimate the potential impact of a vaccine using a transmission model. As there are currently no existing measured estimates of the efficacy of IXCHIQ, we relied on an expert panel consisting of individuals from academia, WHO, CEPI and Gavi to obtain consensus estimates of key characteristics of the vaccine. We used a conservative estimate of 70% protection against disease and 40% protection against infection for the vaccine. The other key characteristics of the vaccine are in Table S5. For endemic settings, we assume that vaccines are introduced to individuals 12 years and over based on current age specific recommendations through an initial campaign followed by supplemental immunisation campaigns, occurring every 5 years. For epidemic settings, we consider the use of a vaccine stockpile where vaccines are distributed in response to an outbreak in a district of 10 million inhabitants, with the timing and size of outbreaks based on our findings above. We assume that there is a delay for the outbreak to be detected (based on a minimum number of cases occurring) and that vaccination occurs over a set duration of time. We assume that transmission is seasonal. To ensure the timings between the reactive campaigns and the start of the outbreak were realistic, we calibrated the simulated outbreak to epidemic time series retrieved from the Brazilian national case notification database (Figure S4). The trajectory of each outbreak, and in particular when the epidemic ends, will depend on contributions of immunity from the vaccines, immunity from infections and seasonality in the force of infection.

We find that on average, achieving 50% vaccination coverage of the population exposed to an outbreak would require 132 million doses per year, (53.9 million in endemic locations and 68.5 million in epidemic locations). The total number of doses is strongly driven by India, where CHIKV circulates endemically (33 million doses per year). We estimate that this level of vaccination, with a vaccine that has 70% efficacy against disease, 40% efficacy against infection and protecting for 5 years, would lead to 4.9 million fewer infections, 651,000 fewer chronic cases, 3,750 fewer deaths and 209,000 DALYs averted per year (Figure 4, Table S3). On average, globally, per 100,000 doses used, we estimate 3,720 (95% CI: 3,170-4,110) infections averted, 2,460 (95% CI: 2,180-2,650) cases averted, 2.84 (95% CI : 2.48-3.08) deaths averted and 158 (95% CI : 138-172) DALYs averted. The impact in epidemic settings was higher than in endemic settings with 4,440 (95% CI : 3,970-4,950) infections averted per 100,000 doses used in epidemic settings compared to 2,680 (95% CI : 1,430-3,290) in endemic settings.

**Figure 4.**
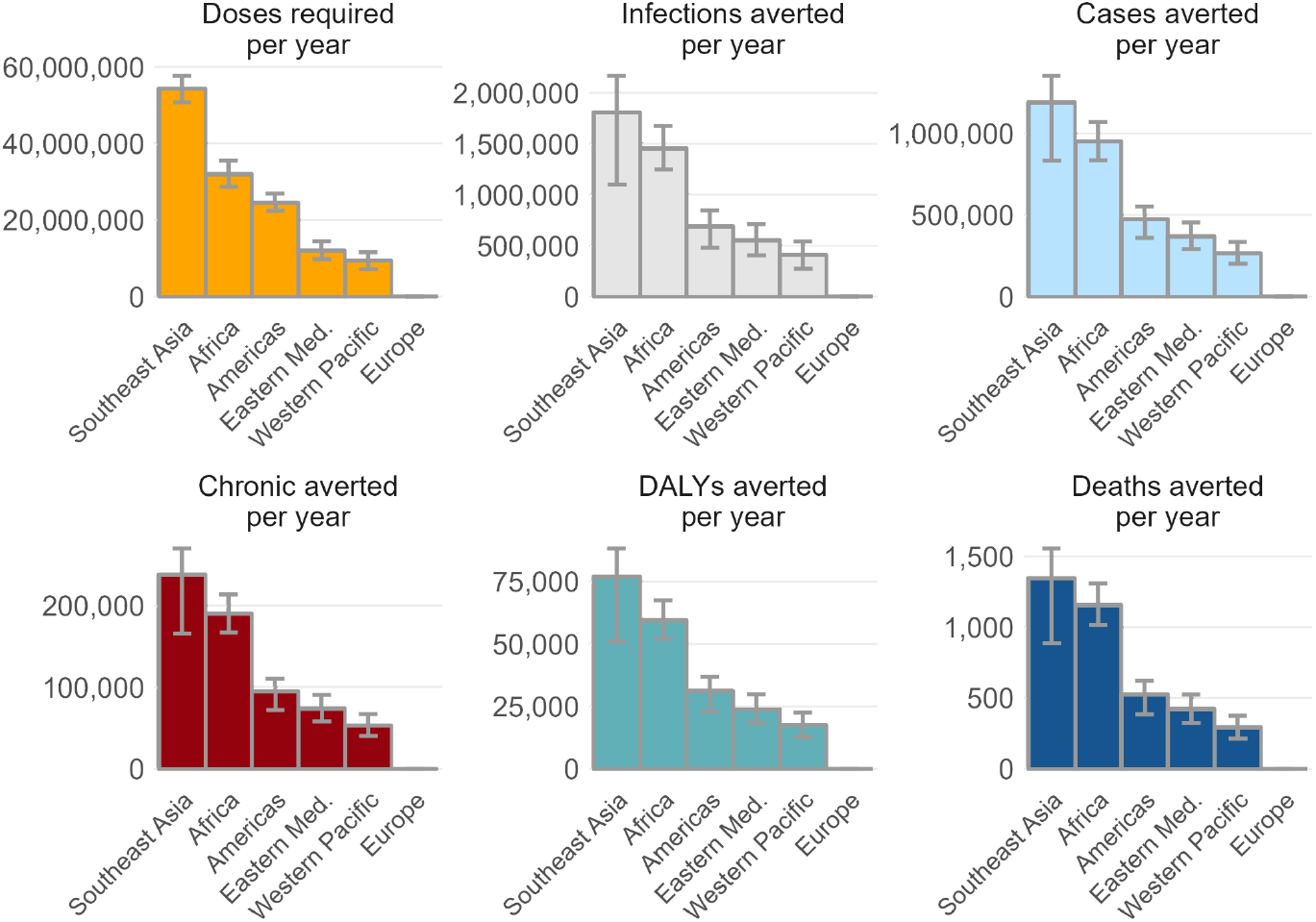
Summary of impact by WHO region for the-base case model

The model results are reliant on a number of strong assumptions linked to vaccine, epidemiological and rollout characteristics. To explore the relative importance of each assumption, we varied each parameter in turn and compared our estimates of the number of infections, cases, deaths and DALYs with those estimated in the base case for epidemic scenarios (Figure 5). We find the conclusions from the model are most sensitive to a range of assumptions about the vaccine characteristics (effectiveness against infection and disease, duration of protection), the rollout strategy (coverage level, time to coverage, time taken to detect outbreaks), the natural history of disease (probability of symptoms). Decreasing the size of subdistricts where a simulated outbreak occurs does not change the average estimates for burden at the country-level (Figure S5).

**Figure 5.**
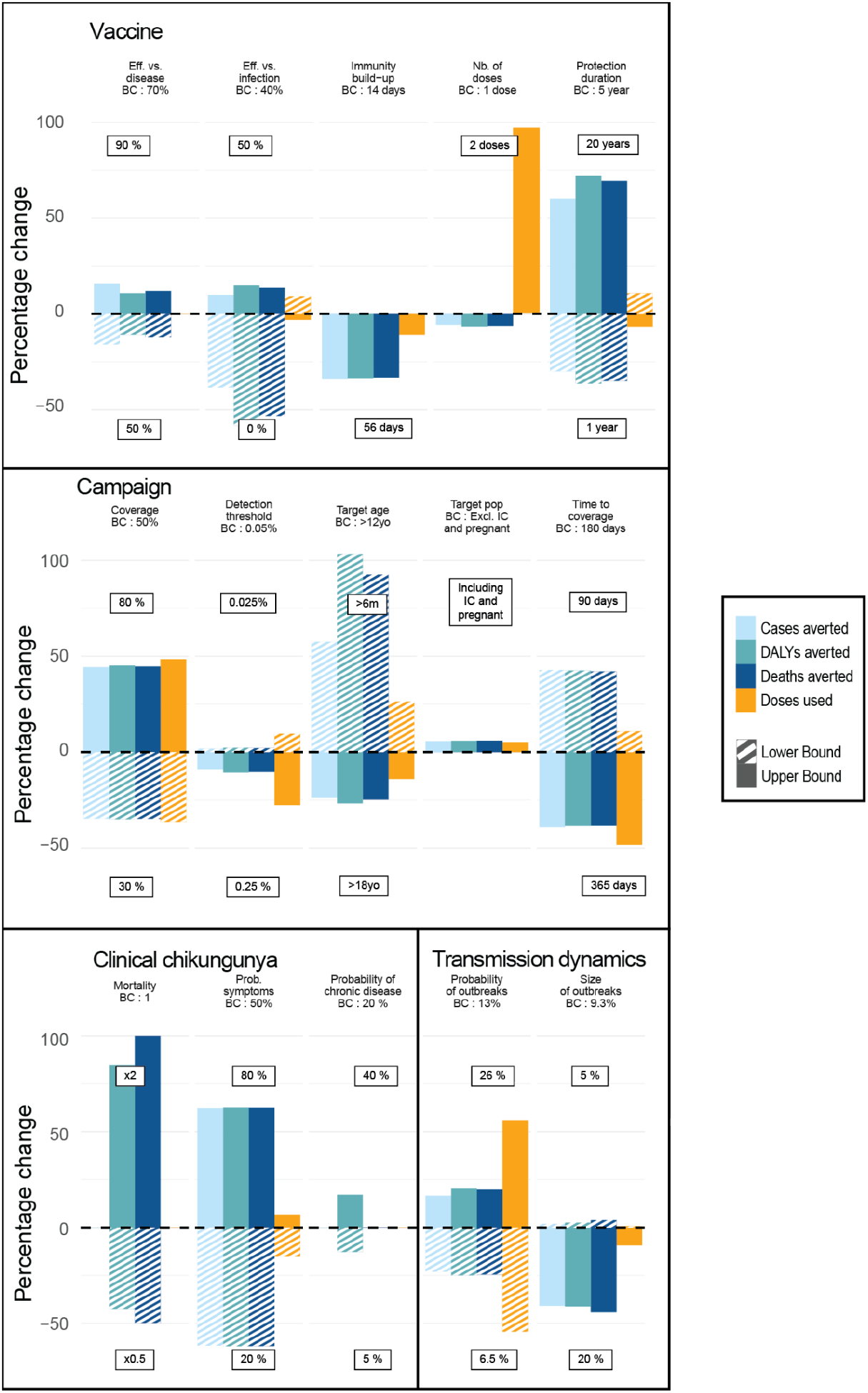
Sensitivity analysis of the model parameters. Percentage change in doses used, cases, DALYs and deaths averted when changing one parameter compared to the base case (BC) scenario.

## Discussion

We have presented a comprehensive overview of the global burden from CHIKV and the potential from the new vaccine. Our findings demonstrate that large portions of the globe are at risk from the pathogen, usually from sporadic outbreaks separated by around 6 years. Our findings suggest that reactive vaccine campaigns using a pre-existing vaccine stockpile could substantially reduce the burden from CHIKV. In heavily endemic areas, such as India and Brazil, routine immunisation would also reduce the impact of the virus on public health.

Tackling CHIKV using the vaccine will require integration of the vaccine into existing immunisation protocols. However, for many parts of the world, this is unlikely to be an attractive prospect if epidemics are infrequent, and the duration of vaccine protection remains poorly understood. In this context, responsive vaccine campaigns from stockpiles are a good alternative, as is done with cholera^11^. However, a stockpile-based approach relies on being able to successfully detect outbreaks. We show that even with a delayed response, where a deployment occurs only once thousands of infections have occurred, the vaccine can still limit the size of the outbreak and the burden from the pathogen. However, with further delays, the potential impact of a vaccine campaign is reduced. Further, in many settings, it has been shown that multiple large CHIKV outbreaks have occurred without a single case reported^4,5^. Improved case detection will be required in many countries, especially in Africa and Asia, to go alongside any stockpile approach.

On average, we estimate that in locations where CHIKV circulates, an average of 1.5% of the susceptible population gets infected each year. These estimates are largely consistent with findings from a previous systematic review that estimated a median force of infection of around 0.7%^10^. Our findings of 19.5 million annual symptomatic cases is around a fifth of the estimated global burden of symptomatic dengue virus (DENV), despite the same areas of the world affected^12^. The differences in CHIKV and DENV burden appear linked to differences in underlying epidemiological patterns, with DENV more capable of transitioning to sustained endemic circulation than chikungunya. The reasons for these inherent differences remain unclear, although are unlikely to be static. The changing climate is leading to larger areas being suitable for *Aedes* vectors, coupled with longer transmission seasons and increased mobility in and out of locations with CHIKV transmission will lead to more frequent CHIKV introductions and sustained outbreaks.

Alongside the long-term chronic sequelae, there is a growing realisation of the deadly nature of the pathogen^13^. We estimate over 20,000 each year lose their lives from CHIKV, far more than actually reported. This difference can be accounted for from the substantial underdetection of outbreaks, and misattribution of cause of death, especially as most CHIKV deaths occur in the elderly who often have other comorbidities^14^. However, we also note that our estimates are dependent on a good understanding of the infection fatality ratio. While the case fatality proportion and how it differs by age, has been reasonably well characterised^2^, the probability that an infected individual becomes a symptomatic and detected case, is less well understood. CHIKV infection is generally considered to result in disease, however, a cohort study in the Philippines with regular blood draws and active disease surveillance found over 80% of infections were subclinical^15–17^. Heterogeneity in the probability of an infected individual becoming a detected case will be driven by differences in healthcare seeking and surveillance systems. There is also the potential that the probability of symptoms differs by CHIKV lineage^18^.

CHIKV vaccines used a correlate of protection as a means to obtain licensure^19^, rather than standard phase III randomised controlled trials. This means there do not exist efficacy estimates that would normally be available. We therefore do not currently know the protection from infection or disease from vaccines or the duration of protection. VLA1553 is a live attenuated vaccine, which generates high titers^20^. It has also been shown that natural infection results in persistent titers, lasting decades, and the presence of any antibody titers was protective against infection and disease^15,19,21^. It may therefore be that our vaccine impact estimates are too low. Assuming an improved profile of 90% protection against infection and disease would lead to 4,560 cases averted per 100,000 doses used compared to 2,460 cases averted in our more conservative assumption of 70% efficacy against disease and 40% against infection. Phase IV trials, which are currently planned for Brazil, will help us understand how realistic these assumptions are.

Dividing CHIKV transmission into ‘epidemic’ and ‘endemic’ countries is a substantial simplification. In particular, it is clear that within countries such as Brazil and India, there will be areas that experience sporadic epidemic transmission, whereas other areas will have more sustained transmission. However, while crude, this division does provide some insight into national patterns of incidence, which are most relevant for developing vaccine strategies. Further, our estimates of the underlying epidemiology of CHIKV and the potential impact of the vaccine are necessarily reliant on strong assumptions. In particular, there is limited understanding about chikungunya transmission, disease history and the efficacy of the new vaccines. We assume that the underlying level of transmission is the same across all epidemic countries, and that transmission levels are the same across endemic countries. While an oversimplification, continent-specific estimates suggest that the dynamics are broadly similar across affected regions (Figure S6). The major exception is South America, which had a higher force of infection than expected. However, CHIKV is new to this continent, and as the virus now encounters substantial population immunity, we can expect the force of infection to fall. Further, we have attempted to account for within-country heterogeneity in the population size at risk of infection by using the relationship between average national estimates of *Aedes* occurrence and history of CHIKV transmission. The relationship between local risk and national average risk is likely to be more complex but is unlikely to result in substantial differences in overall estimates of national burden.

In conclusion, CHIKV is a major threat to public health across much of the globe. However, with new vaccines, we have a real opportunity to combat this threat. Improved abilities to identify and quickly respond to outbreaks will be central to maximising the potential of the vaccine.

## Methods

### Countries and territories considered

We considered countries and territories (as defined by the UN) with a population size of over 200,000 inhabitants. We included territories to avoid problems with some (especially island) territories being in different CHIKV risk zones than the remainder of the country (e.g., French Guiana and mainland France). This resulted in a total of 190 countries and territories.

### Literature review

Taking each country and territory in turn, we used the following resources:

- Google (search term ‘chikungunya AND [COUNTRY]’)
- Google scholar (search term ‘chikungunya AND [COUNTRY]’)
- PubMed (search term ‘chikungunya AND [COUNTRY]’)
- GIDEON
- WHO/PAHO websites
- ProMED (search term ‘chikungunya AND [COUNTRY]’)

We identified Ministry of Health websites where possible. For each country and territory, we attempted to identify whether there had been evidence of CHIKV autochthonous transmission. We considered evidence of infection as either the detection of cases (with at least one case confirmed via PCR or IgM), or from seroprevalence studies (IgG or IgM). For all years prior to and including 2010, we recorded whether there had been evidence of infection. For cases in years 2011-2022, we recorded the specific year where infection had been detected.

### Seroprevalence studies

As part of the literature review process, we specifically highlighted seroprevalence studies for CHIKV for data extraction. Our inclusion criteria were studies in healthy individuals from the general population, which had tested for CHIKV IgG. We excluded seroprevalence studies in suspected cases. From each detected study, we identified the location of the study (from which we subsequently identified coordinates), the number of individuals per age group and the number that tested positive for CHIKV. We subsequently contacted the authors of all identified seroprevalence studies to obtain finer scale data on age and location. From this process, we obtained 40 age-stratified seroprevalence datasets from research groups covering 85 locations across 26 countries (Figure S1).

### Relationship between Aedes mosquito distribution and CHIKV

For each 5 km x 5 km grid cell of a given country, we used the relationship between mosquito occurrence and the probability of CHIKV outbreaks and the number of people living in that cell to obtain a weighted average of the size of the population at risk in that location. For India, China and the United States that have large populations, we added an additional mask that assumes CHIKV transmission only occurs in areas where there is good evidence of sustained transmission, from either national seroprevalence studies (India), or good case reporting (China and the United States)^6^.

We explored the extent to which modelled estimates of *Aedes aegypti* and *Aedes albopictus*, the two vectors for CHIKV, are associated with CHIKV presence in a country. Using estimates of the global distribution of the two vectors and the distribution of the human population distribution per country, we extracted the human population weighted average presence of *Aedes albopictus* and *Aedes aegypti* in each country ^22,23^. We compared these estimates of mosquito presence to the probability of that country having ever reported CHIKV transmission, and found a strong association between the two (Figure 1B).

### Assigning of epidemic status by country

To estimate the reliability of case data for each country, we used the Healthcare Access and Quality (HAQ) Index that is measured on a scale from 0 (worst) to 100 (best) based on amenable mortality ^24^. Countries in the first two deciles of the index (HAQ index >82.2) were classified as having a good surveillance system^24^. We next used the data on case occurrence, seroprevalence studies and mosquito distribution to categorise the epidemic status of all countries and territories. Each country and territory was assigned to ‘endemic’ (i.e., with evidence of sustained transmission each year), ‘epidemic’ (i.e., evidence of transmission but not sustained across years) or ‘no transmission’. We also provide an assessment of the strength of the evidence for the categorization of each country (Table 1).

### Estimating CHIKV transmission dynamics

We used the collected data to inform different models and get estimates on parameters that capture the global dynamics of CHIKV. These models rely on the definition of Force of Infection (FOI) which represents the rate at which the susceptible members of the population in a community get infected. For endemic countries, we estimated a FOI using a serocatalytic model in a Markov chain Monte Carlo (MCMC) framework. In epidemic countries, we considered that FOI was not constant and estimated an annual probability of an outbreak occurring as well as the annual FOI when an outbreak occurs.

We subsetted the serological datasets based on the status classification of the country they originate from. The epidemic-prone countries were used to fit a single epidemic model to estimate an overall probability of outbreak occurrence (µ) and average outbreak size (λ). The endemic countries were used to fit a single endemic model to estimate the distribution of time-constant FOIs across locations characterised by its mean (λ_*endemic*_) and standard deviation (σ_*endemic*_).

### Epidemic model

We assume that outbreaks have a yearly probability µ of occurring with an average size of λ and a standard deviation σ that are the same across locations. In each location, we simulate outbreak patterns and retain the global parameters that generated the most likely patterns. An outbreak pattern in a given location is suitable if it approximates well age-sepcific cumulative forces of infection inferred from the age-stratified seroprevalence in that location.

For a given location r, and a given age group A, we call *N*_*pos,ra*_ the number of samples that tested positive to IgG antibodies against CHIKV and *N* _*tot,rA*_ the total number of samples tested. Note that the size of the age groups differed from one location to another. We use a binomial likelihood to fit our model to the data :

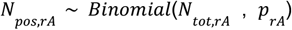

With *p* _*rA*_ being the probability that an individual from a given age group A will have been infected by the time the sample was collected. By definition, the probability *p* of being infected at age a follows an exponential law of rate equal to the FOI (escaping infection from birth to age a). And the *p* _*rA*_ (proportion of people that have been infected from birth to age a) is the cumulative distribution function of *p*. Taking into account grouping by age, we get :

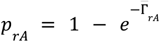

With 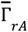 being the average cumulative FOI for an age group A that we estimated to be the mean value of the cumulative FOI Γ_*rN*_ for all ages included in the year group :

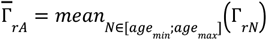

From their birth to the study year, any individual that is N years old will have been exposed to the cumulative FOI :

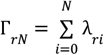

With λ_*ri*_ being the annual FOI in location r at year (*Y* ‒ *i*) where *Y* is the year where the serum samples were collected in this location.

Ultimately, λ_*ri*_ are simulated during an iterated filtering process drawn from *Beta*(α, β) with a probability µ and set to 0 otherwise. α and β are calculated such that the mean and standard deviation of the distribution are equal to λ and σ. For parameter identifiability purposes, σ was fixed at 0.0025. The parameters estimated are µ and λ using a Sequential Monte Carlo (SMC) Bayesian framework embedded in C and accessed using R and the *pomp* ^*25*^ package. Uninformative uniform priors *U*(0, 1) were used for λ and σ. 1,000 particles divided into equal-sized blocks (one by location) were used to simulate outbreak patterns and explore the parameter space. Each particle filtering instance was replicated 20 times to estimate the likelihood and associated uncertainty.

### Endemic model

Following the previous notations, we now assume that outbreaks occur every year and aim to estimate the average force of infection resulting from that endemic pattern. We assume all the λ_*ri*_ to be equal to λ _*r*_ such that

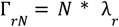

Where λ_*r*_ is the time-constant FOI estimated for location *r*. λ_*r*_ are drawn from a beta distribution *Beta*(α, β) of mean λ_*endemic*_ and standard deviation σ_*endemic*_. The hyperparameters λ_*endemic*_ and σ_*endemic*_ are also estimated in our model using uninformative uniform priors *U*(0, 1) for both. We used a Markov Chain Monte Carlo (MCMC) Bayesian framework using Stan and R. The posterior distribution of considered parameters was sampled using 4 chains of 4,500 iterations each including a burn-in phase of 500 iterations.

### Vaccine simulation framework

For each country, we simulate transmission of chikungunya. If a country has more than 10 million inhabitants, we considered it was structured with several 10-million population subdistricts with independent FOI patterns from one to another.

The population has an age structure derived from the 2020 UN World Population Prospects ^26^. They were grouped in the 12 following age groups : 0-5, 6-10, 11-12, 13-18, 19-20, 21-30, 31-40, 41-50, 51-60, 61-70, 71-80, 81+. We use these age groups to allow for age-specific vaccine policies (especially where we consider 12+ and 18+ vaccination strategies), as well as allowing sufficient granularity to have age-specific mortality.

Each age group has six different compartments that individuals can be allocated to depending on their infection and vaccination status :

- S : unvaccinated, never-infected individuals
- I : unvaccinated, infectious individuals
- R : unvaccinated, recovered individuals (seropositives)
- V : vaccinated never-infected individuals
- IV : vaccinated, infectious individuals
- RV : vaccinated, recovered individuals (seropositives)

We run the simulation for a period of 20 years during which we measure the impact estimates.

Prior to the first year, the FOIs for each year are drawn. If the country is endemic, a time-constant FOI is drawn from a Beta distribution of mean λ_*endemic*_ and standard deviation σ_*endemic*_. If the country is epidemic, for each year, the annual FOI is set to 0 (no transmission event) with probability (1 ‒ µ), and to a location specific FOI drawn from a Beta distribution of mean λ_*endemic*_ and standard deviation σ_*endemic*_ with probability µ.

Every year the following events occur :

- Loss of vaccination protection
- Running a deterministic SIRV model with seasonal transmissibility calculated from the FOI previously drawn.
- Ageing of the population

### Loss of vaccination

The proportion of vaccinated people in the population exponentially decays with a decay rate 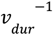 where ν_*dur*_ (base case 5 years) is the vaccine duration of protection. This is translated by a flow of individuals from compartment V to S and from compartment RV to R.

### Vaccination schedule

In endemic countries, the vaccination campaign occurs every 5 years and vaccinates 50% of the unvaccinated population (S+R) over 180 days. Vaccinations occur at a fixed daily rate over the duration of the vaccination campaign.

In epidemic countries, vaccination starts as soon as a threshold number of cases is detected (500 cases per million), and also aims at vaccinating 50% of the unvaccinated population (S+R) over 180 days. However, vaccination can stop prematurely if the outbreak dies out reaching 50 daily cases per million.

We assume the vaccine to be delivered in a single dose, with a time to reach protective immunity of 14 days. The vaccine provides a protection against infection of 40% and protection against disease of 70%.

### SIRV model

The chikungunya transmission and vaccination of the population are simulated simultaneously using an age-stratified SIRV model described by the following system of differential equations for a given age group a (Figures S6-S7):

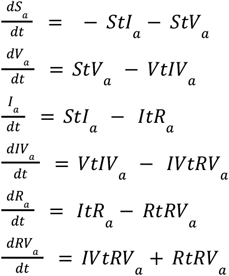

With :

- 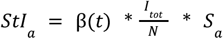
- 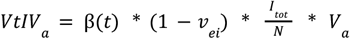
- *ItR*_*a*_ = σ * *I*_*a*_
- *IVtRV*_*a*_ = σ * *IV*_*a*_
- *StV*_*a*_ and *RtRV*_*a*_ being dictated by the daily rates of vaccination as described previously
- 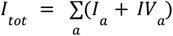, the total number of infected individuals.
- 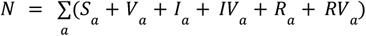, the total population size
- ν_*ei*_ the vaccine induced protection against infection
- 1/σ is the mean duration of infectiousness
- β(*t*) is the seasonal transmission rate, linearly decreasing over time (sawtooth pattern with an offsetted baseline)

### Population ageing

Each year, individuals from compartments of age *a* go to compartments of age *a+1*. A new birth cohort of completely susceptible individuals is introduced in the population at age 0 and individuals in the last age compartment (100+) are removed from the population. In order to keep the age structure constant through time, each compartment is then proportionally adjusted.

### Model parameters

As there is considerable uncertainty in the vaccine characteristics and feasibility of different deployment strategies, there was a meeting of experts convened, with representatives from WHO, CEPI, Gavi and academia, where a broad consensus was reached. These are summarised in Tables S4, S5.

Unvaccinated infections have a 50% chance of being symptomatic (expert opinion) and vaccinated infections have 50% * (1-vaccine protection against disease)/(1-vaccine protection against infection) = 50% * (1-0.7)/(1-0.4) = 25% chance of developing symptoms. We assume that all symptomatic infections are detected by the surveillance systems (number of cases). (Figure S9)

Symptomatic infections have a 50 % chance of having a mild acute phase and 50% of having a severe one ^27,28^. Severe acute phases have then a 40% chance of developing chronic symptoms (arthralgia) ^29–32^. We assume the acute phases to last for 7 days on average and chronic symptoms to last for 1 year^1,33,34^.

The probability of death given symptomatic infection (CFR) is age-dependant and was based from Brazilian clinical data ^2^

Years Lived with a Disability (YLDs) were computed by assuming a disability weight of 0.006 for mild acute symptoms, 0.133 for severe acute symptoms and 0.233 for arthralgia (chronic symptoms). ^35^ Years of Life Lost (YLLs) were measured as being the difference between the age of death and the mean life expectancy of the country. Individuals dying after the mean life expectancy of the country did not contribute to the YLL calculation.

By definition, Disability-adjusted life years lost to chikungunya (DALYs) were calculated as being the sum of YLLs and YLDs.

## Data Availability

The publicly available seroprevalence datasets will be made available on GitHub, along with the references of their article of origin where relevant.

https://github.com/G-ribeiro-dos-santos/chik-global-burden

## Supplementary materials

**Table S1.** Number of countries and territories by epidemic status by WHO region.

|  | Endemic transmission |  | Epidemic transmission |  | No transmission |  |
| --- | --- | --- | --- | --- | --- | --- |
|  | Good evidence | Poor evidence | Good evidence | Poor evidence | Good evidence | Poor evidence |
| Africa | 2 | 2 | 14 | 25 | 1 | 2 |
| The Americas | 1 | 0 | 29 | 0 | 1 | 0 |
| Eastern Mediterranean | 0 | 0 | 7 | 2 | 5 | 7 |
| Europe | 0 | 0 | 2 | 0 | 34 | 15 |
| Southeast Asia | 1 | 0 | 8 | 1 | 0 | 1 |
| Western Pacific | 0 | 0 | 10 | 0 | 7 | 3 |
| **Total** | **4** | **2** | **70** | **28** | **48** | **28** |

**Table S2.** Summary of annual burden by WHO region.

| WHO Region | Classification | Infections | Cases | Deaths | DALYs | Chronic cases |
| --- | --- | --- | --- | --- | --- | --- |
| Africa | Endemic | 1,880,000<br>(95% CI : 620,000-3,880,000) | 941,000<br>(95% CI : 310,000-1,940,000) | 1,280 (95% CI : 418-2,640) | 67,200 (95% CI : 21,900-139,000) | 188,000 (95% CI : 62,000-388,000) |
| Africa | Epidemic | 5,960,000<br>(95% CI : 2,370,000-11,000,000) | 2,980,000<br>(95% CI : 1,190,000-5,510,000) | 4,080 (95% CI : 1,620-7,570) | 196,000 (95% CI : 78,300-360,000) | 596,000 (95% CI : 237,000-1,100,000) |
| Americas | Endemic | 2,850,000<br>(95% CI : 205,000-8,620,000) | 1,420,000<br>(95% CI : 103,000-4,310,000) | 1,930 (95% CI : 139-5,830) | 83,500 (95% CI : 6,030-253,000) | 285,000 (95% CI : 20,500-862,000) |
| Americas | Epidemic | 2,150,000<br>(95% CI : 344,000-4,870,000) | 1,070,000<br>(95% CI : 172,000-2,430,000) | 1,490 (95% CI : 235-3,370) | 67,000 (95% CI : 10,700-151,000) | 215,000 (95% CI : 34,400-487,000) |
| Eastern Mediterranean | Endemic | - | - | - | - | - |
| Eastern Mediterranean | Epidemic | 3,780,000<br>(95% CI : 602,000-9,580,000) | 1,890,000<br>(95% CI : 301,000-4,790,000) | 2,510 (95% CI : 396-6,370) | 129,000 (95% CI : 20,600-327,000) | 378,000 (95% CI : 60,200-958,000) |
| Europe | Endemic | - | - | - | - | - |
| Europe | Epidemic | 108.00 (95% CI : 0-330.00) | 54.00 (95% CI : 0-165.00) | 0.00 (95% CI : 0-0.00) | 3.14 (95% CI : 0-9.57) | 11.00 (95% CI : 0-33.00) |
| Southeast Asia | Endemic | 9,070,000<br>(95% CI : 660,000-27,900,000) | 4,530,000<br>(95% CI : 330,000-13,900,000) | 5,930 (95% CI : 432-18,200) | 275,000 (95% CI : 20,000-847,000) | 907,000 (95% CI : 66,000-2,790,000) |
| Southeast Asia | Epidemic | 5,430,000<br>(95% CI :<br>1,350,000-12,200,000) | 2,720,000<br>(95% CI :<br>674,000-6,120,000) | 3,620 (95%<br>CI :<br>894-8,130) | 163,000 (95%<br>CI :<br>40,600-368,000) | 543,000 (95% CI :<br>135,000-1,220,000) |
| WesternPacifc | Endemic | - | - | - | - | - |
| WesternPacifc | Epidemic | 2,600,000<br>(95% CI :<br>254,000-6,890,000) | 1,300,000<br>(95% CI :<br>127,000-3,450,000) | 1,730 (95%<br>CI :<br>167-4,590) | 84,500 (95% CI :<br>8,140-226,000) | 260,000 (95% CI :<br>25,400-689,000) |
| Global | Endemic | 13,800,000<br>(95% CI :<br>3,910,000-33,100,000) | 6,900,000<br>(95% CI :<br>1,950,000-16,500,000) | 9,140 (95%<br>CI :<br>2,610-21,800) | 426,000 (95%<br>CI :<br>125,000-1,010,000) | 1,380,000 (95% CI :<br>391,000-3,310,000) |
| Global | Epidemic | 19,900,000<br>(95% CI :<br>11,800,000-30,600,000) | 9,960,000<br>(95% CI :<br>5,900,000-15,300,000) | 13,400 (95%<br>CI :<br>7,960-20,600) | 640,000 (95%<br>CI :<br>381,000-982,000) | 1,990,000 (95% CI :<br>1,180,000-3,060,000) |
| Global | Endemic and epidemic | 33,700,000<br>(95% CI :<br>19,800,000-55,500,000) | 16,900,000<br>(95% CI :<br>9,910,000-27,800,000) | 22,600 (95%<br>CI :<br>13,300-36,900) | 1,070,000<br>(95% CI :<br>634,000-1,740,000) | 3,370,000 (95% CI :<br>1,980,000-5,550,000) |

**Table S3.** Summary of annual impact by WHO region for the base case model.

| WHO region | classification | Doses required | Infections averted | Cases averted | Deaths averted | DALYs averted | Chronic cases averted |
| --- | --- | --- | --- | --- | --- | --- | --- |
| Africa | Endemic | 6,620,000<br>(95% CI : 6,620,000-6,620,000) | 208,000 (95% CI : 121,000-292,000) | 132,000 (95% CI : 88,000-173,000) | 169 (95% CI : 108-227) | 9,380 (95% CI : 6,000-12,500) | 26,300 (95% CI : 17,600-34,600) |
| Africa | Epidemic | 25,400,000<br>(95% CI : 22,100,000-28,900,000) | 1,250,000 (95% CI : 1,040,000-1,450,000) | 819,000 (95% CI : 705,000-930,000) | 988 (95% CI : 844-1,130) | 50,100 (95% CI : 42,700-57,400) | 164,000 (95% CI : 141,000-186,000) |
| Americas | Endemic | 14,200,000<br>(95% CI : 14,200,000-14,200,000) | 304,000 (95% CI : 98,100-404,000) | 219,000 (95% CI : 111,000-257,000) | 243 (95% CI : 109-298) | 14,700 (95% CI : 6,780-17,600) | 43,800 (95% CI : 22,200-51,500) |
| Americas | Epidemic | 10,300,000<br>(95% CI : 8,210,000-12,700,000) | 386,000 (95% CI : 263,000-510,000) | 256,000 (95% CI : 190,000-318,000) | 284 (95% CI : 206-359) | 16,500 (95% CI : 11,900-21,000) | 51,200 (95% CI : 38,100-63,600) |
| EasternMed | Endemic | - | - | - | - | - | - |
| EasternMed | Epidemic | 12,000,000<br>(95% CI : 9,790,000-14,500,000) | 555,000 (95% CI : 406,000-709,000) | 370,000 (95% CI : 291,000-453,000) | 424 (95% CI : 324-526) | 24,100 (95% CI : 18,200-29,900) | 74,100 (95% CI : 58,200-90,600) |
| Europe | Endemic | - | - | - | - | - | - |
| Europe | Epidemic | 0 (95% CI : 0-0) | 0 (95% CI : 0-0) | 0 (95% CI : 0-0) | 0 (95% CI : 0-0) | 0 (95% CI : 0-0) | 0 (95% CI : 0-0) |
| SoutheastAsia | Endemic | 33,100,000<br>(95% CI : 33,100,000-33,100,000) | 931,000 (95% CI : 263,000-1,210,000) | 617,000 (95% CI : 290,000-723,000) | 718 (95% CI : 289-872) | 41,400 (95% CI : 17,300-49,000) | 123,000 (95% CI : 58,100-145,000) |
| SoutheastAsia | Epidemic | 21,200,000<br>(95% CI :<br>17,800,000-24,500,000) | 878,000 (95%<br>CI :<br>671,000-1,070<br>,000) | 573,000<br>(95% CI :<br>468,000-670,<br>000) | 629 (95%<br>CI :<br>503-743) | 35,700<br>(95% CI :<br>28,500-42,<br>200) | 115,000<br>(95% CI :<br>93,500-134,<br>000) |
| WesternPacific | Endemic | - | - | - | - | - | - |
| WesternPacific | Epidemic | 9,390,000<br>(95% CI :<br>7,260,000-1<br>1,600,000) | 410,000 (95%<br>CI :<br>279,000-542,0<br>00) | 266,000<br>(95% CI :<br>203,000-334,<br>000) | 295 (95%<br>CI :<br>217-375) | 17,600<br>(95% CI :<br>12,900-22,<br>400) | 53,200<br>(95% CI :<br>40,500-66,8<br>00) |
| **Global** | Endemic | 53,900,000<br>(95% CI :<br>53,900,000-53,900,000) | 1,440,000<br>(95% CI :<br>771,000-1,770<br>,000) | 968,000<br>(95% CI :<br>629,000-1,11<br>0,000) | 1,130 (95%<br>CI :<br>692-1,330) | 65,500<br>(95% CI :<br>40,700-75,<br>500) | 194,000<br>(95% CI :<br>126,000-22<br>1,000) |
| **Global** | Epidemic | 78,400,000<br>(95% CI :<br>72,100,000-84,500,000) | 3,480,000<br>(95% CI :<br>3,110,000-3,8<br>50,000) | 2,280,000<br>(95% CI :<br>2,100,000-2,<br>480,000) | 2,620 (95%<br>CI :<br>2,390-2,860<br>) | 144,000<br>(95% CI :<br>131,000-1<br>58,000) | 457,000<br>(95% CI :<br>420,000-49<br>6,000) |
| **Global** | **Endemic**<br>**and**<br>**epidemic** | **132,000,00**<br>**0 (95% CI :**<br>**126,000,00**<br>**0-138,000,0**<br>**00)** | **4,920,000**<br>**(95% CI :**<br>**4,220,000-5,4**<br>**40,000)** | **3,250,000**<br>**(95% CI :**<br>**2,890,000-3,**<br>**510,000)** | **3,750 (95%**<br>**CI :**<br>**3,290-4,080**<br>**)** | **209,000**<br>**(95% CI :**<br>**184,000-2**<br>**28,000)** | **650,000**<br>**(95% CI :**<br>**577,000-70**<br>**1,000)** |

**Table S4.** Model parameters for CHIKV epidemiology and natural history of disease and ranges used for sensitivity analysis

| Name | Base case | Range for sensitivity | Detail |
| --- | --- | --- | --- |
| **CHIKV transmission - epidemic settings** |  |  |  |
| Frequency of outbreaks | 0.13 per year | 0.07-0.27 | Estimated from seroprevalence data |
| Average outbreak size | 9.4% | 5-20% | Estimated from seroprevalence data |
| Standard deviation in outbreak size | 0.05 | - | Estimated from seroprevalence data |
| Baseline immunity | WHO region specific | - | Estimated from seroprevalence data |
| **CHIKV transmission - endemic settings** |  |  |  |
| Mean force of infection | 3.1% | - | Estimated from seroprevalence data |
| Standard deviation in force of infection | 0.019 | - | Estimated from seroprevalence data |
| Baseline immunity | WHO region specific | 0-0.5 | Estimated from seroprevalence data |
| **Natural history of disease** |  |  |  |
| Duration of infectiousness | 7 days | - | 36-38 |
| Prob. symptoms | 0.5 | 0.2-0.8 | Expert opinion |
| Case fatality proportion | Age dependent (mean ~0.001) | - | 2 |
| DALY per infection | Age dependent (mean ~0.082) | - | 35,39,40 |

**Table S5.**
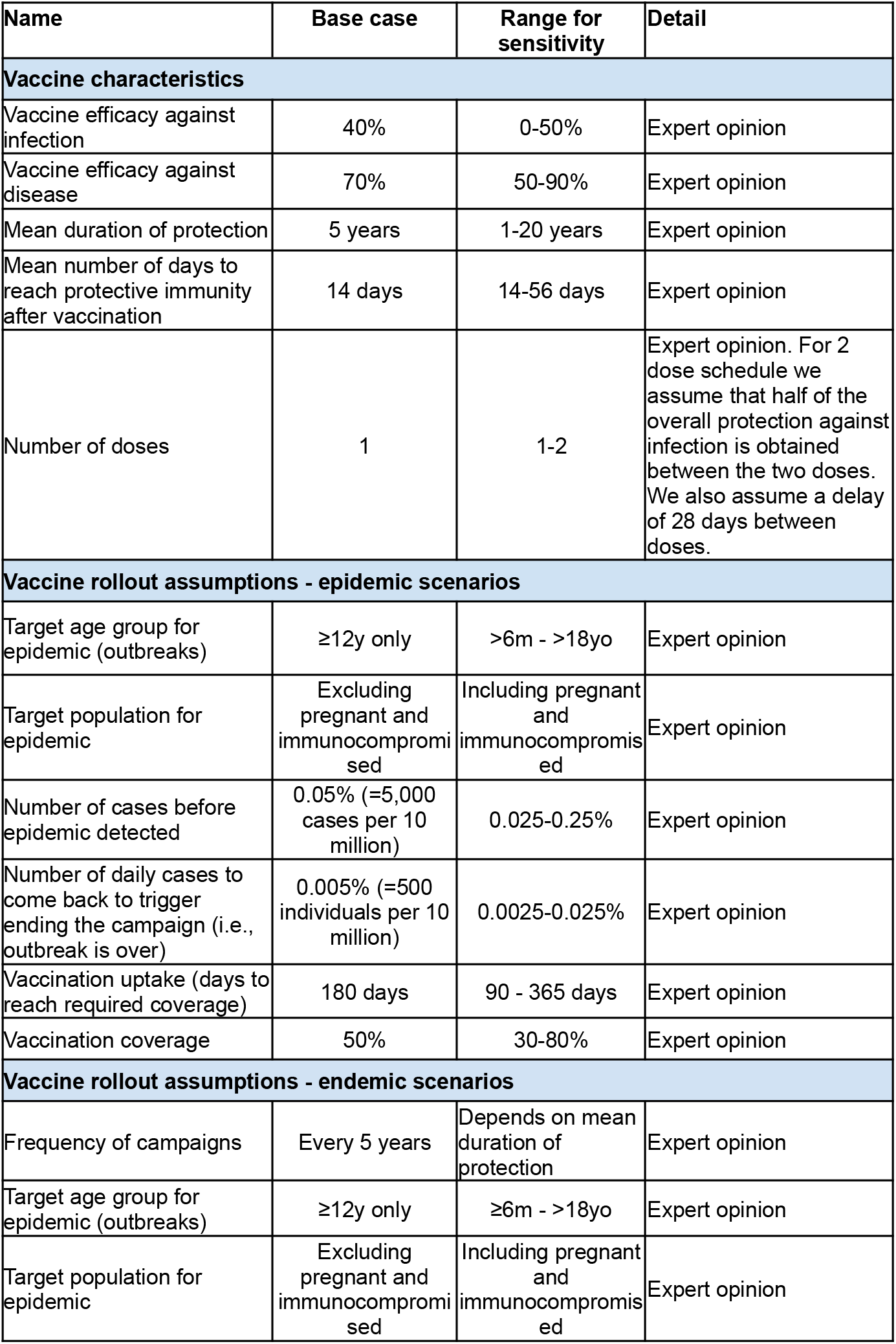
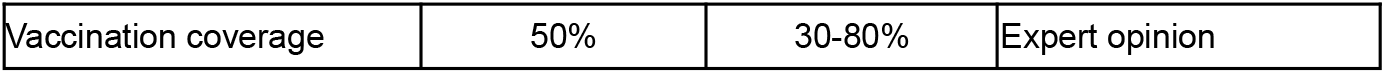
Model parameters for vaccine characteristics and rollout assumptions and ranges used for sensitivity analysis

| Name | Base case | Range for sensitivity | Detail |
| --- | --- | --- | --- |
| **Vaccine characteristics** |  |  |  |
| Vaccine efficacy against infection | 40% | 0-50% | Expert opinion |
| Vaccine efficacy against disease | 70% | 50-90% | Expert opinion |
| Mean duration of protection | 5 years | 1-20 years | Expert opinion |
| Mean number of days to reach protective immunity after vaccination | 14 days | 14-56 days | Expert opinion |
| Number of doses | 1 | 1-2 | Expert opinion. For 2 dose schedule we assume that half of the overall protection against infection is obtained between the two doses. We also assume a delay of 28 days between doses. |
| **Vaccine rollout assumptions - epidemic scenarios** |  |  |  |
| Target age group for epidemic (outbreaks) | ≥12y only | >6m - >18yo | Expert opinion |
| Target population for epidemic | Excluding pregnant and immunocompromised | Including pregnant and immunocompromised | Expert opinion |
| Number of cases before epidemic detected | 0.05% (=5,000 cases per 10 million) | 0.025-0.25% | Expert opinion |
| Number of daily cases to come back to trigger ending the campaign (i.e., outbreak is over) | 0.005% (=500 individuals per 10 million) | 0.0025-0.025% | Expert opinion |
| Vaccination uptake (days to reach required coverage) | 180 days | 90 - 365 days | Expert opinion |
| Vaccination coverage | 50% | 30-80% | Expert opinion |
| **Vaccine rollout assumptions - endemic scenarios** |  |  |  |
| Frequency of campaigns | Every 5 years | Depends on mean duration of protection | Expert opinion |
| Target age group for epidemic (outbreaks) | ≥12y only | ≥6m - >18yo | Expert opinion |
| Target population for epidemic | Excluding pregnant and immunocompromised | Including pregnant and immunocompromised | Expert opinion |
| Vaccination coverage | 50% | 30-80% | Expert opinion |

**Table S6.** Average impact per dose

| **Classification** | **Infections averted per 100,000 doses** | **Cases averted per 100,000 doses** | **Deaths averted per 100,000 doses** | **DALYs averted per 100,000 doses** | **Chronic cases averted per 100,000 doses** |
| --- | --- | --- | --- | --- | --- |
| Endemic | 2,680 (95% CI : 1,430-3,290) | 1,800 (95% CI : 1,170-2,050) | 2.10 (95% CI : 1.28-2.46) | 121 (95% CI : 75.5-140) | 359 (95% CI : 233-410) |
| Epidemic | 4,440 (95% CI : 3,970-4,950) | 2,920 (95% CI : 2,650-3,200) | 3.35 (95% CI : 3.03-3.68) | 184 (95% CI : 166.0-203) | 584 (95% CI : 531-640) |
| Endemic and Epidemic | 3,720 (95% CI : 3,170-4,110) | 2,460 (95% CI : 2,180-2,650) | 2.84 (95% CI : 2.48-3.08) | 158 (95% CI : 138.0-172) | 492 (95% CI : 436-531) |

**Figure S1.**
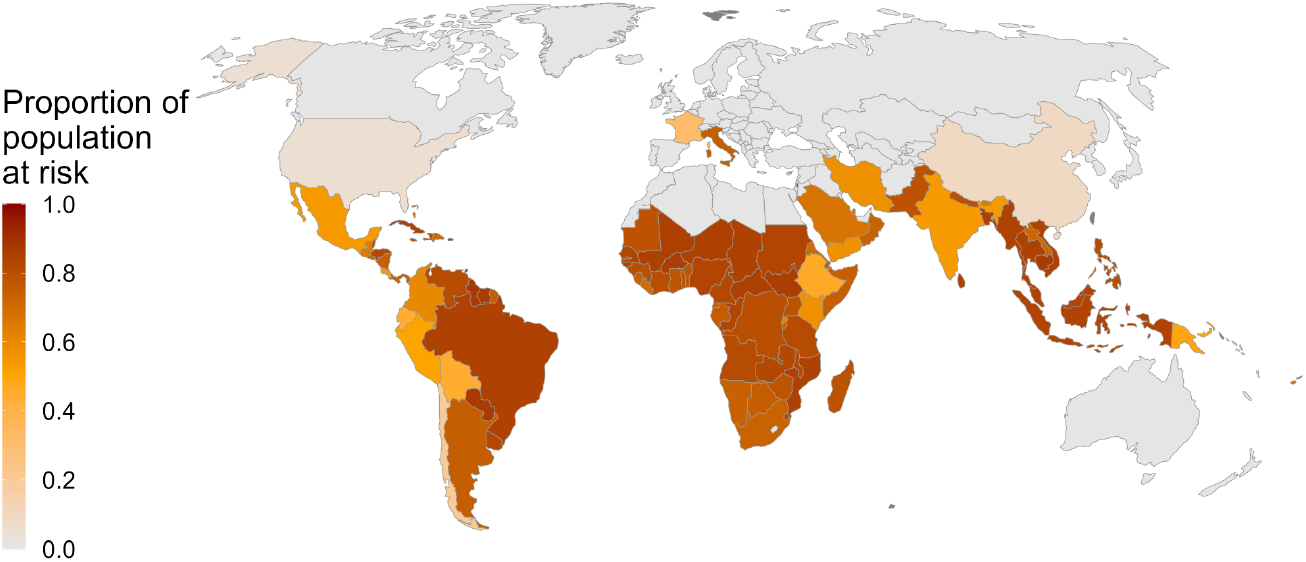
Proportion of population at risk per country

**Figure S2.**
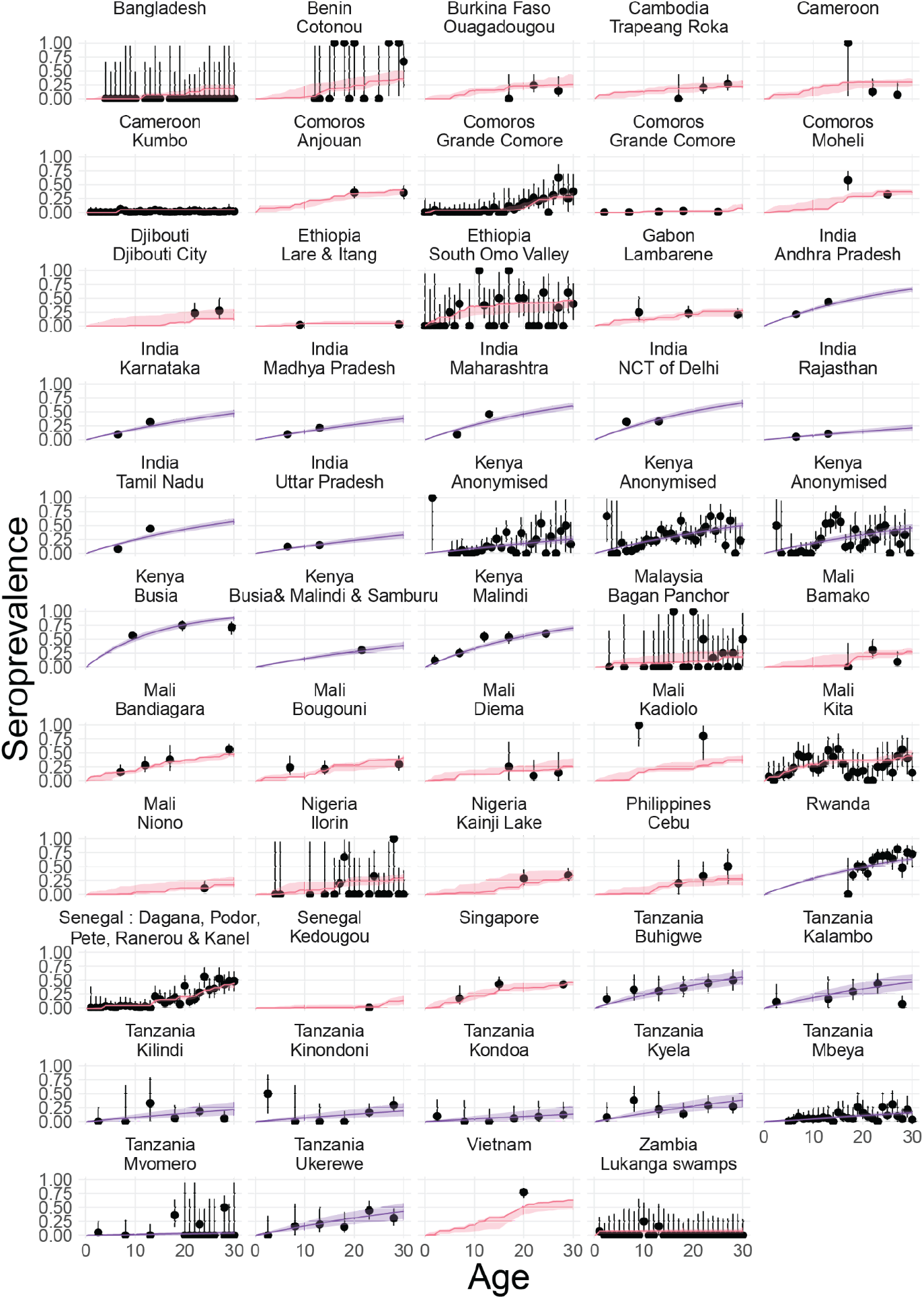
Fits of serocatalytic model. Seroprevalence estimates and 95%CI in black. Epidemic model fits in red. Endemic model fits in blue.

**Figure S3.**
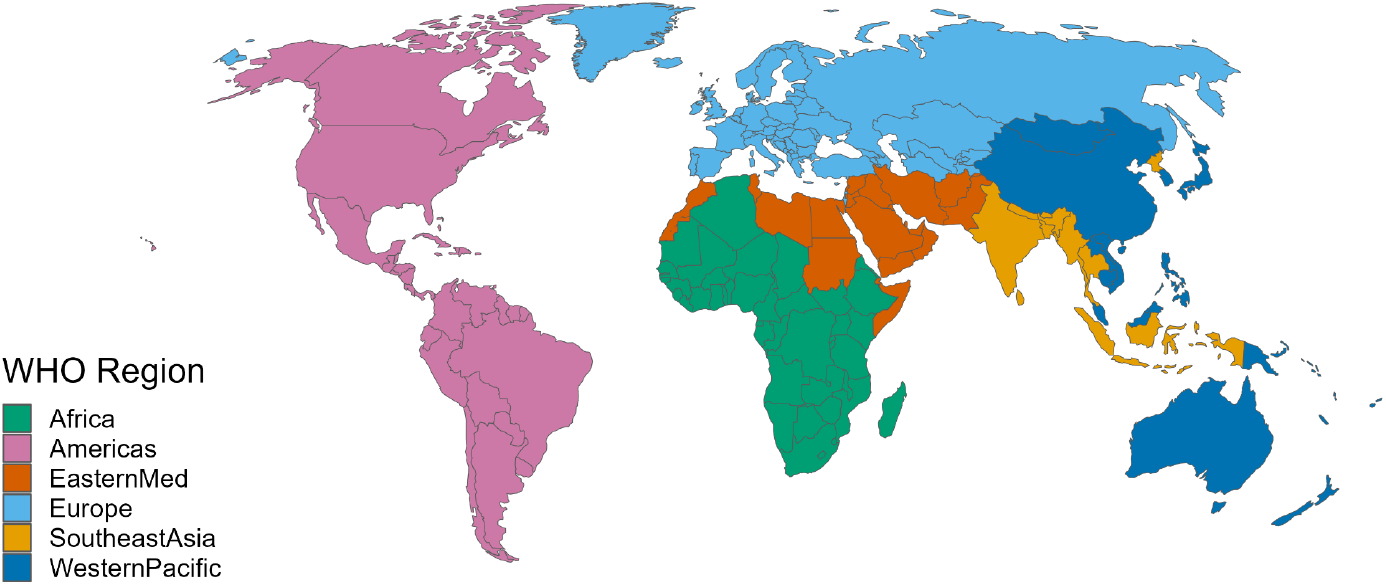
WHO region per country.

**Figure S4.**
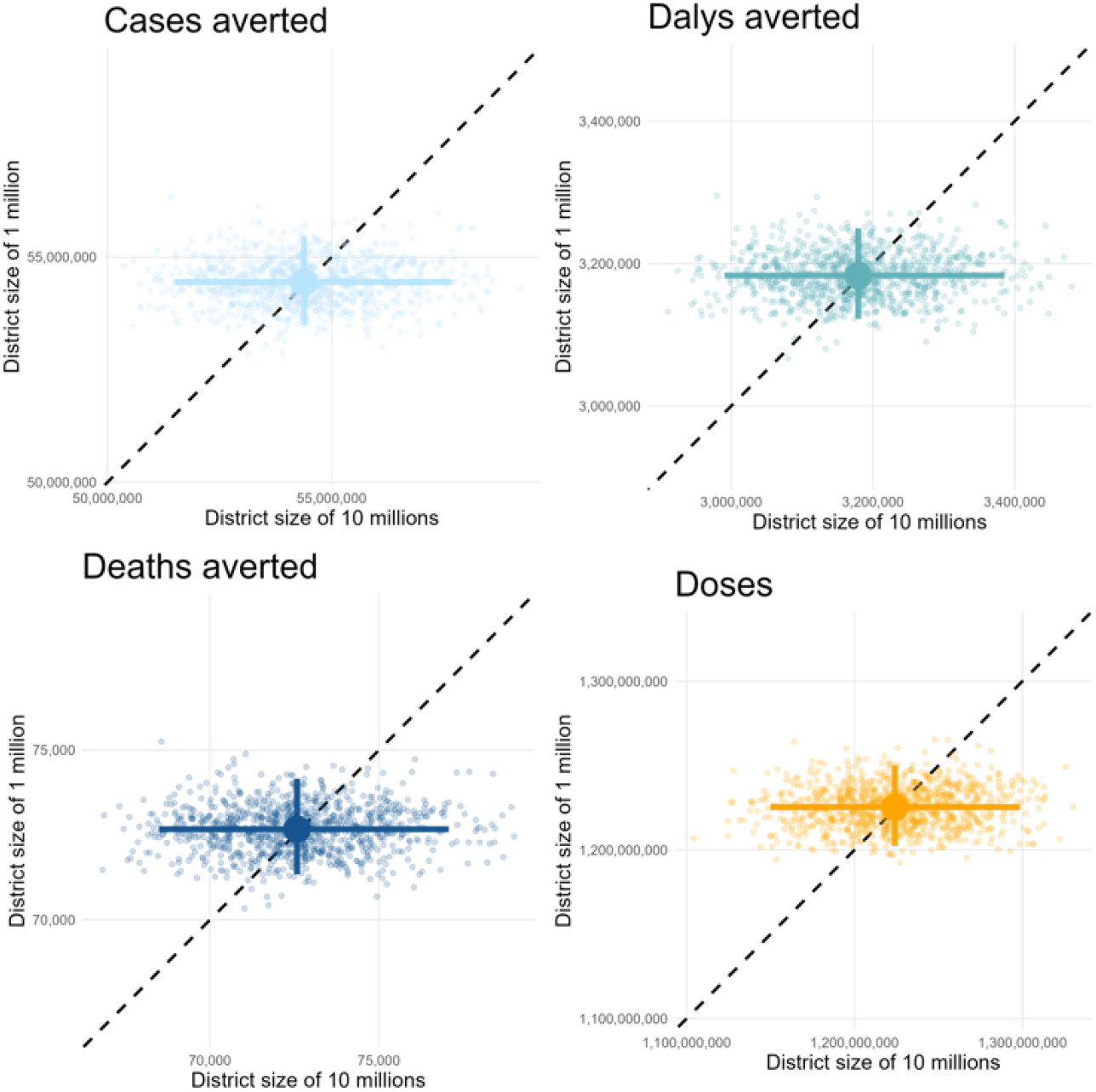
Sensitivity analysis on the impact of the population size on the districts where the simulated outbreak occurs.

**Figure S5.**
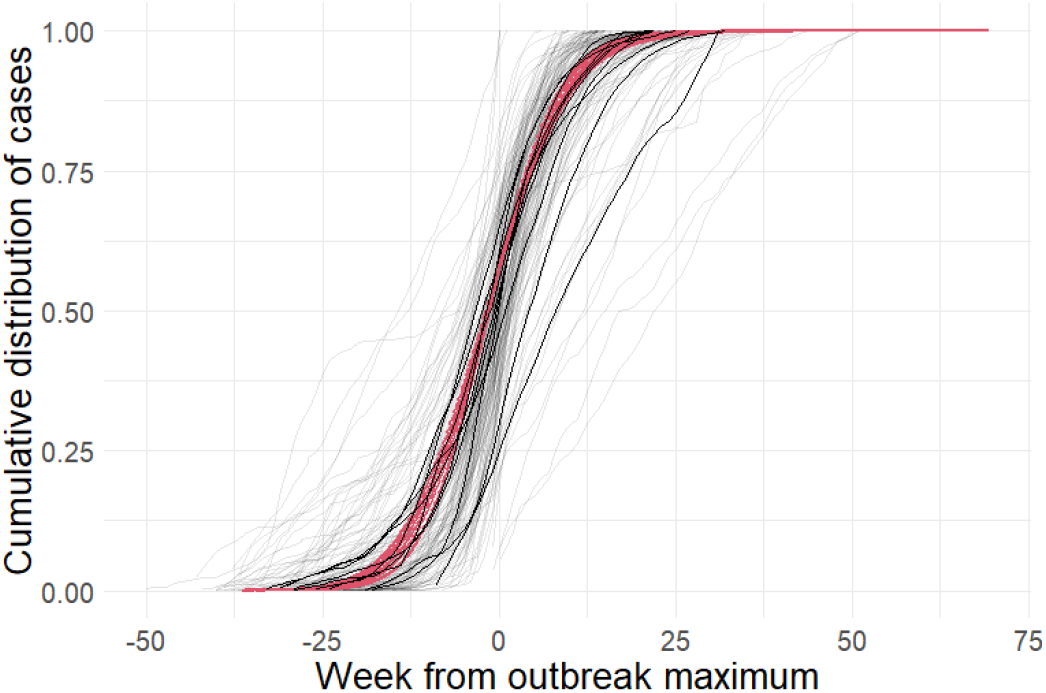
Shape and timing of observed outbreaks against simulated outbreaks. Empirical distribution function of chikungunya cases reported in Brazil since 2015 by state (thin black lines) and nationally (plain black lines). Empirical distribution function of simulated outbreaks with varying force of infection (red lines).

**Figure S6.**
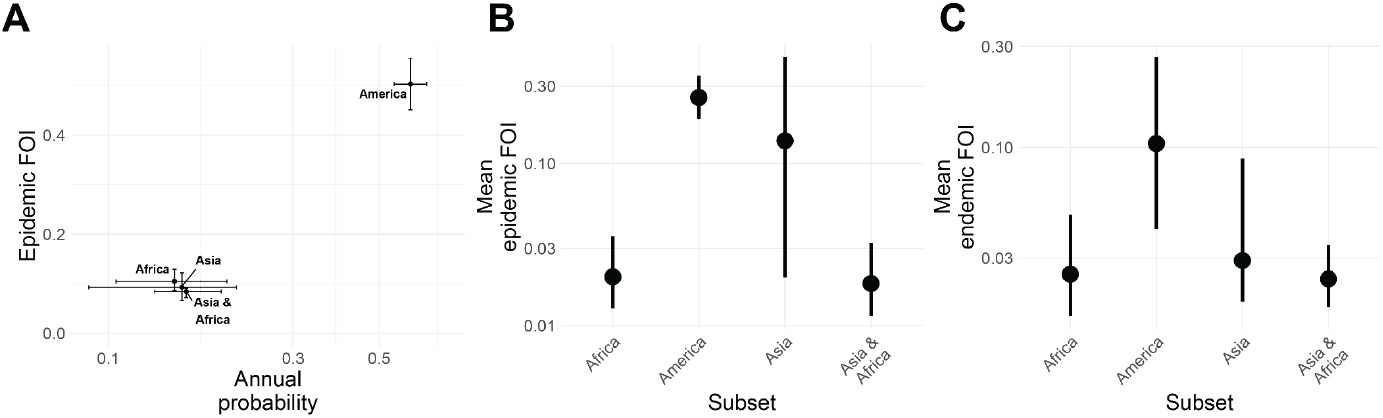
Continent-specific estimates for transmission dynamics. Estimates and 95%CI for different subset of continents from **(A)** the epidemic serocatalytic model **(B)** the endemic serocatalytic model fitted on data form epidemic countries **©** the endemic serocatalytic model fitted on data from endemic countries

**Figure S7.**
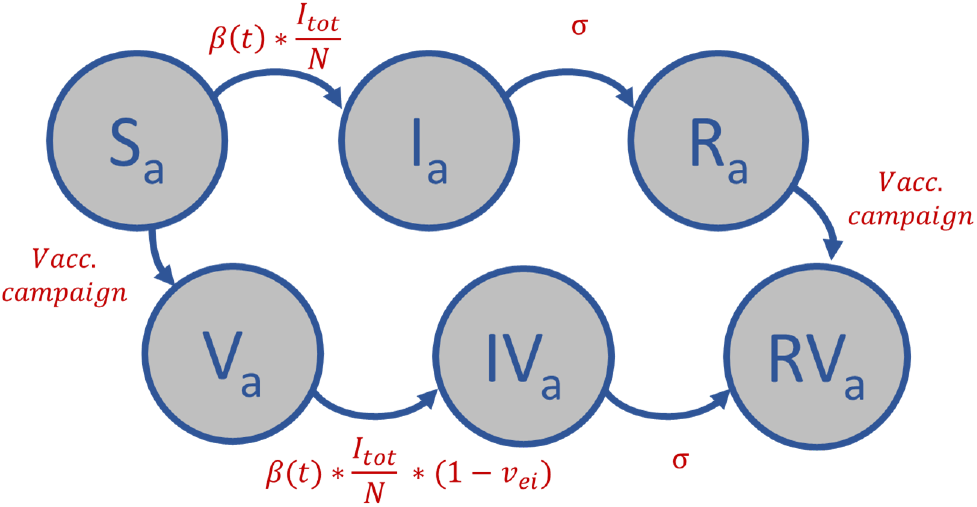
Structure of the SIRV model for a given age group.

**Figure S8.**
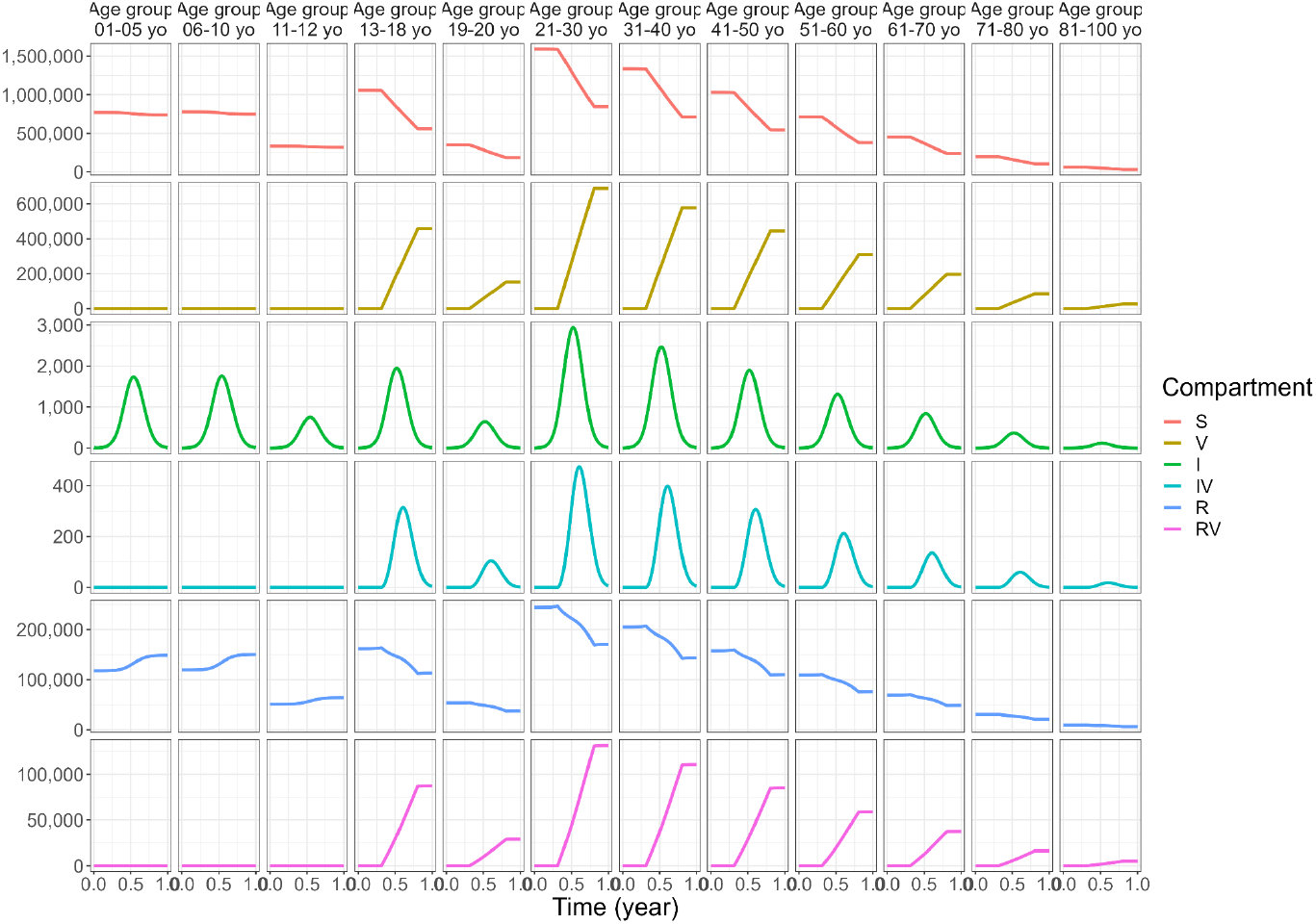
Outbreak simulated for the base case vaccination scenario over one year in a representative population of 10 million people.

**Figure S9.**
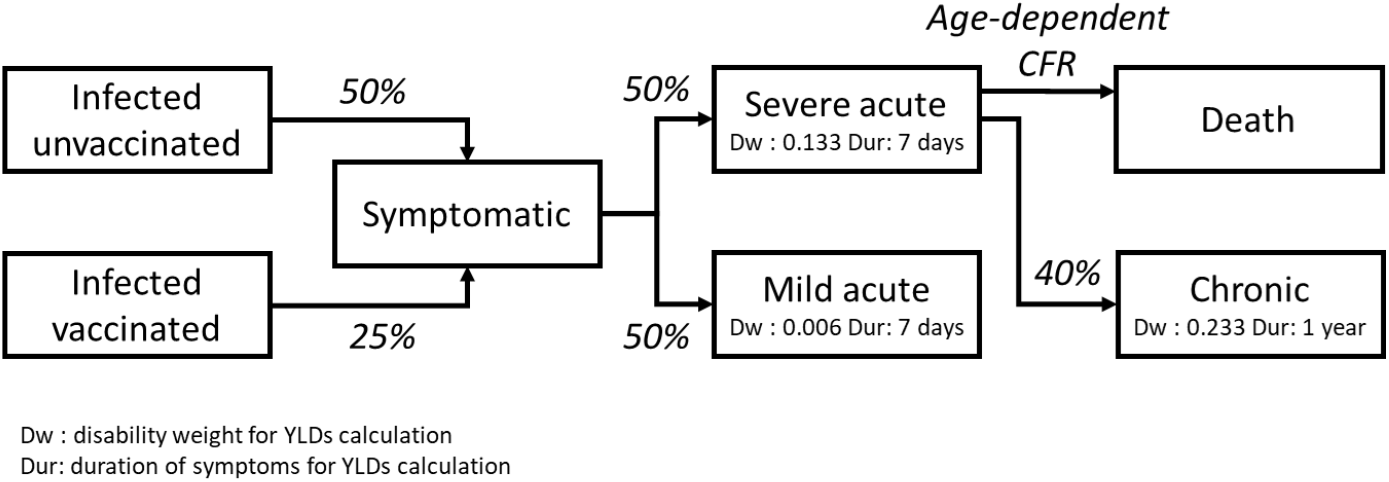
Summary diagram for YLDs calculation

## Inclusion and ethics statement

*All models were developed using anonymized datasets. The data was either extracted from existing publications or provided by the underlying data collectors without any personal identifiers. The datasets are credited to the local research groups that generated them. Local and regional research has been duly acknowledged in citations*.

## Code availability data

*All the code used for the analysis presented here is publicly available in the following Github repository G-ribeiro-dos-santos/chik-global-burden*.

## References

1. O’Driscoll, M., Salje, H., Chang, A. Y. & Watson, H. Arthralgia resolution rate following chikungunya virus infection. Int. J. Infect. Dis. 112, 1–7 (2021).

2. de Souza, W. M. et al. Spatiotemporal dynamics and recurrence of chikungunya virus in Brazil: an epidemiological study. Lancet Microbe 4, e319–e329 (2023).

3. The overlapping global distribution of dengue, chikungunya, Zika and yellow fever.

4. Salje, H. et al. Reconstruction of 60 Years of Chikungunya Epidemiology in the Philippines Demonstrates Episodic and Focal Transmission. J. Infect. Dis. 213, 604–610 (2016).

5. Kyungah Lim, J. et al. Seroepidemiological reconstruction of long-term chikungunya virus circulation in Burkina Faso and Gabon. J. Infect. Dis. jiac246 (2022).

6. Kumar, M. S. et al. Seroprevalence of chikungunya virus infection in India, 2017: a cross-sectional population-based serosurvey. Lancet Microbe 2, e41–e47 (2021).

7. Office of the Commissioner. FDA Approves First Vaccine to Prevent Disease Caused by Chikungunya Virus. U.S. Food and Drug Administration https://www.fda.gov/news-events/press-announcements/fda-approves-first-vaccine-prevent-disease-caused-chikungunya-virus (2023).

8. More than US$ 1.8 billion in support for African vaccine manufacturing, catching up missed children and pandemic preparedness approved as Gavi Board steps up efforts to tackle backsliding and fight health emergencies. https://www.gavi.org/news/media-room/initiatives-african-vaccine-manufacturing-approved-gavi-board (2023).

9. Godaert, L. et al. Misdiagnosis of Chikungunya Virus Infection: Comparison of Old and Younger Adults. J. Am. Geriatr. Soc. 66, 1768–1772 (2018).

10. Kang, H. et al. Chikungunya Seroprevalence, Force of Infection, and Prevalence of Chronic Disability in Endemic and Epidemic Settings: Systematic Review, Meta-Analysis, and Modelling Study. (2023) doi:10.2139/ssrn.4617040.

11. Desai, S. N. et al. Achievements and challenges for the use of killed oral cholera vaccines in the global stockpile era. Hum. Vaccin. Immunother. 13, 579–587 (2017).

12. Messina, J. P. et al. A global compendium of human dengue virus occurrence. Sci Data 1, 140004 (2014).

13. Salje, H. & Cortés Azuero, O. The deadly potential of chikungunya virus. Lancet Infect. Dis. 24, 442–444 (2024).

14. Cerqueira-Silva, J. M. P. Risk of death following chikungunya virus disease the 100 Million Brazilian Cohort, 2015-2018: A matched cohort study and self-controlled case series. Lancet Infect. Dis. (2024).

15. Yoon, I.-K. et al. High rate of subclinical chikungunya virus infection and association of neutralizing antibody with protection in a prospective cohort in the Philippines. PLoS Negl. Trop. Dis. 9, e0003764 (2015).

16. Hennessey, M. J. et al. Seroprevalence and Symptomatic Attack Rate of Chikungunya Virus Infection, United States Virgin Islands, 2014-2015. Am. J. Trop. Med. Hyg. 99, 1321–1326 (2018).

17. Sissoko, D. et al. Seroprevalence and risk factors of chikungunya virus infection in Mayotte, Indian Ocean, 2005-2006: a population-based survey. PLoS One 3, e3066 (2008).

18. Bustos Carrillo, F. et al. Epidemiological evidence for lineage-specific differences in the risk of inapparent Chikungunya virus infection. J. Virol. 93, (2019).

19. Milligan, G. N., Schnierle, B. S., McAuley, A. J. & Beasley, D. W. C. Defining a correlate of protection for chikungunya virus vaccines. Vaccine 37, 7427–7436 (2019).

20. Valneva reports positive 12-month Antibody persistence data for single-shot Chikungunya vaccine candidate. Valneva https://valneva.com/press-release/valneva-reports-positive-12-month-antibody-persistence-data-for-single-shot-chikungunya-vaccine-candidate/ (2022).

21. Yoon, I.-K. et al. Pre-existing chikungunya virus neutralizing antibodies correlate with risk of symptomatic infection and subclinical seroconversion in a Philippine cohort. Int. J. Infect. Dis. 95, 167–173 (2020).

22. Kraemer, M. U. G. et al. The global distribution of the arbovirus vectors Aedes aegypti and Ae. albopictus. Elife 4, e08347 (2015).

23. Bright, E., Coleman, P. & Dobson, J. E. LandScan: A global population database for estimating populations at risk. Photogramm. Eng. Remote Sens. 66, 849–858 (2000).

24. Fullman, N. et al. Measuring performance on the Healthcare Access and Quality Index for 195 countries and territories and selected subnational locations: a systematic analysis from the Global Burden of Disease Study 2016. Lancet 391, 2236–2271 (2018).

25. King, A. A., Nguyen, D. & Ionides, E. L. Statistical Inference for Partially Observed Markov Processes via the R Package pomp. arXiv [stat.ME] (2015).

26. World Population Prospects - Population Division - United Nations. https://population.un.org/wpp/.

27. Man, O. M., Fuller, T. L., Rosser, J. I. & Nielsen-Saines, K. Re-emergence of arbovirus diseases in the State of Rio de Janeiro, Brazil: The role of simultaneous viral circulation between 2014 and 2019. One Health 15, 100427 (2022).

28. Bautista-Reyes, E., Núñez-Avellaneda, D., Alonso-Palomares, L. A. & Salazar, M. I. Chikungunya: Molecular Aspects, Clinical Outcomes and Pathogenesis. Rev. Invest. Clin. 69, 299–307 (2017).

29. Morens, D. M. & Fauci, A. S. Chikungunya at the door--déjà vu all over again? N. Engl. J. Med. 371, 885–887 (2014).

30. Chang, A. Y. et al. Frequency of Chronic Joint Pain Following Chikungunya Virus Infection: A Colombian Cohort Study. Arthritis Rheumatol 70, 578–584 (2018).

31. Cardona-Ospina, J. A., Rodriguez-Morales, A. J. & Villamil-Gómez, W. E. The burden of Chikungunya in one coastal department of Colombia (Sucre): Estimates of the disability adjusted life years (DALY) lost in the 2014 epidemic. J. Infect. Public Health 8, 644–646 (2015).

32. Puntasecca, C. J., King, C. H. & LaBeaud, A. D. Measuring the global burden of chikungunya and Zika viruses: A systematic review. PLoS Negl. Trop. Dis. 15, e0009055 (2021).

33. Feldstein, L. R. et al. Persistent Arthralgia Associated with Chikungunya Virus Outbreak, US Virgin Islands, December 2014-February 2016. Emerg. Infect. Dis. 23, 673–676 (2017).

34. Warnes, C. M. et al. Longitudinal Analysis of the Burden of Post-Acute Chikungunya-Associated Arthralgia in Children and Adults: A Prospective Cohort Study in Managua, Nicaragua (2014-2019). medRxiv (2023) doi:10.1101/2023.05.09.23289726.

35. Salomon, J. A. et al. Disability weights for the Global Burden of Disease 2013 study. Lancet Glob Health 3, e712–23 (2015).

36. Ruiz-Moreno, D., Vargas, I. S., Olson, K. E. & Harrington, L. C. Modeling dynamic introduction of Chikungunya virus in the United States. PLoS Negl. Trop. Dis. 6, e1918 (2012).

37. Báez-Hernández, N., Casas-Martínez, M., Danis-Lozano, R. & Velasco-Hernández, J. X. A mathematical model for Dengue and Chikungunya in Mexico. bioRxiv 122556 (2017) doi:10.1101/122556.

38. Pialoux, G., Gaüzère, B.-A., Jauréguiberry, S. & Strobel, M. Chikungunya, an epidemic arbovirosis. Lancet Infect. Dis. 7, 319–327 (2007).

39. Yaseen, H. M., Simon, F., Deparis, X. & Marimoutou, C. Estimation of lasting impact of a Chikungunya outbreak in Reunion Island. Epidemiology: Open Access 2, 1–6 (2012).

40. Vidal, E. R. N., Frutuoso, L. C. V., Duarte, E. C. & Peixoto, H. M. Epidemiological burden of Chikungunya fever in Brazil, 2016 and 2017. Trop. Med. Int. Health 27, 174–184 (2022).

